# Mass-Standardised IgG Response to Fourteen SARS-CoV-2 Spike Protein variants and Antibody Subclass analysis for IgG subclasses and IgE for a Long COVID Patient Cohort

**DOI:** 10.64898/2026.01.26.26344863

**Authors:** Abishek Elangovan, Daniel Harper, Philip H. James-Pemberton, Shivali Kohli, Ciara E. Watson, Andrew M. Shaw

## Abstract

A multivariant total subclass analysis has been performed for a control cohort (*n*=15) and a long COVID patient cohort (*n*=15) measuring the IgG1, IgG2, IgG3, IgG4 and IgE response to the following 14 variants of SARS-CoV-2: Wuhan, Alpha, Delta, BA.1, BA.2, BA.5, EG.5.1, XBB.1.5, BA.2.75, CH.1.1, BA.2.12.1, BQ.1.1, JN.1, and KP.3. Significant differences (*p* < 0.05 and *p* < 0.005) between concentrations of IgG subclasses by variant were found in 24% of variants and in mean-normalised distributions. The medians of the mean-normalised distributions were significantly lower for IgG1 (*p* < 0.05) in long COVID patients compared with controls, and significantly higher (*p* < 0.005) for levels of IgG3, IgG4 and IgE for long COVID patients. A preliminary diagnostics classification analysis performed by variation of the mean-normalised upper and lower percentiles symmetrically for IgG3 showed a long COVID diagnostic sensitivity of 80%, and specificity of 80% for the 60^th^ percentile threshold of the control cohort. Three types of long COVID can be identified: patients with at least one variant below the threshold (hypo-immune), patients with at least one variant above the threshold (hyperimmune) and patients with IgG3 levels within the reference range. The multivariant subclass spectrum indicates IgG4 and IgE elevations due to potential chronic antigen exposure from persistent virus or autoimmunity and may indicate potential therapeutic interventions.

## Introduction

Antibody class switching is a measure of the inflammatory status within the body^1^. It occurs when CD4^+^ T cells recognise an antigen presented by a naïve B-cell; the T-cell then releases cytokines to activate the B-cell, enabling differentiation into antibody-secreting plasma cells. The specific cytokine signals arising from CD4^+^ T cells and antigen presenting B-cell interactions, and CD40/CD40L co-stimulation^2-4^, determine the specific antibody subclass, although IgG4 production is more complex^5^. A subclass analysis, including IgE, provides insight into health and disease states, especially chronic inflammatory conditions^6^. Furthermore, elevated IgG4 has been observed in autoimmune conditions^7,8^ such as Crohn’s Disease and ME/CFS^9^.

The relative abundance of each subclass in healthy human sera is IgG1 65 %, IgG2 25 %, IgG3 6 % and IgG4 4 % ^10^. Median IgG1 and IgG3 levels have been reported to rise in response to an influenza vaccine, increasing from 435 and 167 ng/mL pre vaccination to 4422 and 2020 ng/mL 42 days post vaccination^11^, respectively. These changes in IgG1 and IgG3 subtypes are also produced in response to viral infections, and in response to protein-based antigens e.g. SARS-CoV-2 S-protein ^12,13^. Conversely, IgG2 dominates in bacterial infections^1^ although disease severity following SARS-CoV-2 and influenza infections have been associated with IgG2 subclass switching^14-17^. Further, previous studies have shown that mRNA vaccines can induce a high proportion of spike-specific IgG4 responses^18^. Antibody isotype class analysis has not been extensively studied in long COVID patients although mechanisms of viral persistence with sporadic reactivation^19^ and autoimmunity or hyperimmunity^20,21^ have been proposed. Enhanced IgG4 class switching has been observed in individuals with two or more mRNA vaccinations to SARS-CoV-2, and the abundance of IgG4 could be used as a marker for clinical improvement in patients suffering from long COVID^22^.

Long COVID remains a generalised term for more than more than 250 symptoms that occur as post-acute sequalae to SARS-CoV-2 (PASC) exposure and possibly even vaccination^23^. Persistent viral particles and mRNA^24-26^ have been found in tissue biopsies^27^, and circulating spike protein^28-30^ has been reported in 60% of long COVID patients^28^, suggesting viral reactivation mechanisms^19^. Viral reservoirs may be present due to the poor clearance of viral particles by antibodies produced in response to SARS-CoV-2 infection. In this form, the virus is undetectable by immune cells and therefore continues to persist and can disseminate ^31^. Some case studies indicate that these reservoirs can reactivate replication in response to immunosuppressive events, leading to inflammation and infection events throughout the body ^32^. Cases of autoimmunity^33-35^ suggest a characteristic antibody response, potentially including IgG4, as seen in other autoimmune conditions^7,8^ such as Crohn’s Disease. The complexity of the underlying symptoms with a reported eight longitudinal profiles^36^ suggest an evolving IgG subclass distribution and evolving IgE response.

In this study, the total, mass-standardised, IgG antibody concentration and quality (avidity) was measured simultaneously, to multiple SARS-CoV-2 spike proteins: Wuhan, Alpha, Delta, BA.1, BA.2, BA.5, BA.2.75, BA.2.12.1, CH.1.1, BQ.1.1, EG.5.1, XBB.1.5, JN.1 and KP.3. Then, in five separate experiments, the concentration of IgG subclasses: IgG1, IgG2, IgG3 and IgG4 were measured, along with IgE. A retrospective cohort of control, vaccinated patients (*n* = 15) from the pandemic are compared with a cohort of long COVID patients with symptoms persistent for more than 20 weeks, (*n*=15). The distributions for each variant were determined and Mann-Whitney U tests were performed for significance in medians. Mean-normalised subclass distributions were combined for all variants and tested for variation significance (F-test). The correlation between antibody concentration and quality was determined for each subclass, deriving both correlation coefficients and significance testing the slopes (Fisher r-to-*z* transformation).

## Materials and methods

### Materials

Patient IgG and IgE concentration was determined using a multiplexed biophotonic array, measuring the light scattered from the biophotonic surface using an array reader (LISCAR), which uses surface plasmon resonance to detect antibody binding events, described in detail elsewhere^37^. Briefly, this technology utilises a localised gold nanoparticle plasmon sensor which scatters light proportional to the mass of material, formally known as the refractive index, within the plasmon field of the nanoparticle surface. The gold nanoparticles were functionalised with 14 SARS-CoV-2 variants onto two sensor arrays across two LISCAR-6 instruments (Figure 2a). Two SARS-CoV-2 variants were common on the sensor arrays (Wuhan and KP.3) to allow for comparison of coefficient of variation (CV) measurements and for equivalency between the two array readings.

The spike proteins of SARS-CoV-2 variants measured within array one (LC1) were Wuhan, Alpha, Delta, BA.1, BA.2, BA.5, EG.5.1, and KP.3. The variants on array two (LC2) were Wuhan, XBB.1.5, BA.2.75, CH.1.1, BA.2.12.1, BQ.1.1, JN.1, and KP.3. Moreover, three additional proteins were common on both arrays to measure other parameters: recombinant Human Serum Albumin (rHSA) as a reference channel to correct for non-specific binding and detecting bulk changes in the refractive index to ensure the signal is due to antibody-antigen interactions, anti-C-reactive protein (CRP) as a marker of inflammation, and protein A/G as a measure of total IgG.

IgG1 spike reference materials from Sinobiological (40590-D001) were used to calibrate the surface density of spike protein for each variant, binding to the conserved S2 region. No reference material existed for IgG2, IgG3, IgG4 and IgE to spike protein. The detection antibodies for IgG1-4 are all monoclonal antibodies so the ratios between IgG1-4 are all consistent: the IgE detection antibody is polyclonal so there is an unknown, fixed stochiometric binding factor associated, likely a factor ≤ 2, depending on the steric hindrance of binding to one or other binding epitope (Table S1). Consequently, IgG1 is a fully calibrated mg/L response whereas all other channels are all in mass-standardised unit (msu)/L.

### Patient samples

Two patient cohorts were tested; the control cohort (CC, *n* = 15) consisting of samples collected from non-long COVID individuals with no reported symptoms of post-viral sequalae and no known, active SARS-CoV-2 infection at the time of sampling, Table 1, whilst the long COVID cohort (LC, *n* = 15) is comprised of long COVID patients under the care of their own clinician. Patient samples were prepared to a 1:20 concentration using Attomarker buffer, RB1, before being vortexed and filtered using a 0.2 µM filter [Cytiva; 6765-1302] to remove fats from the sample mixture, prior to running the assay.

**Table 1.**
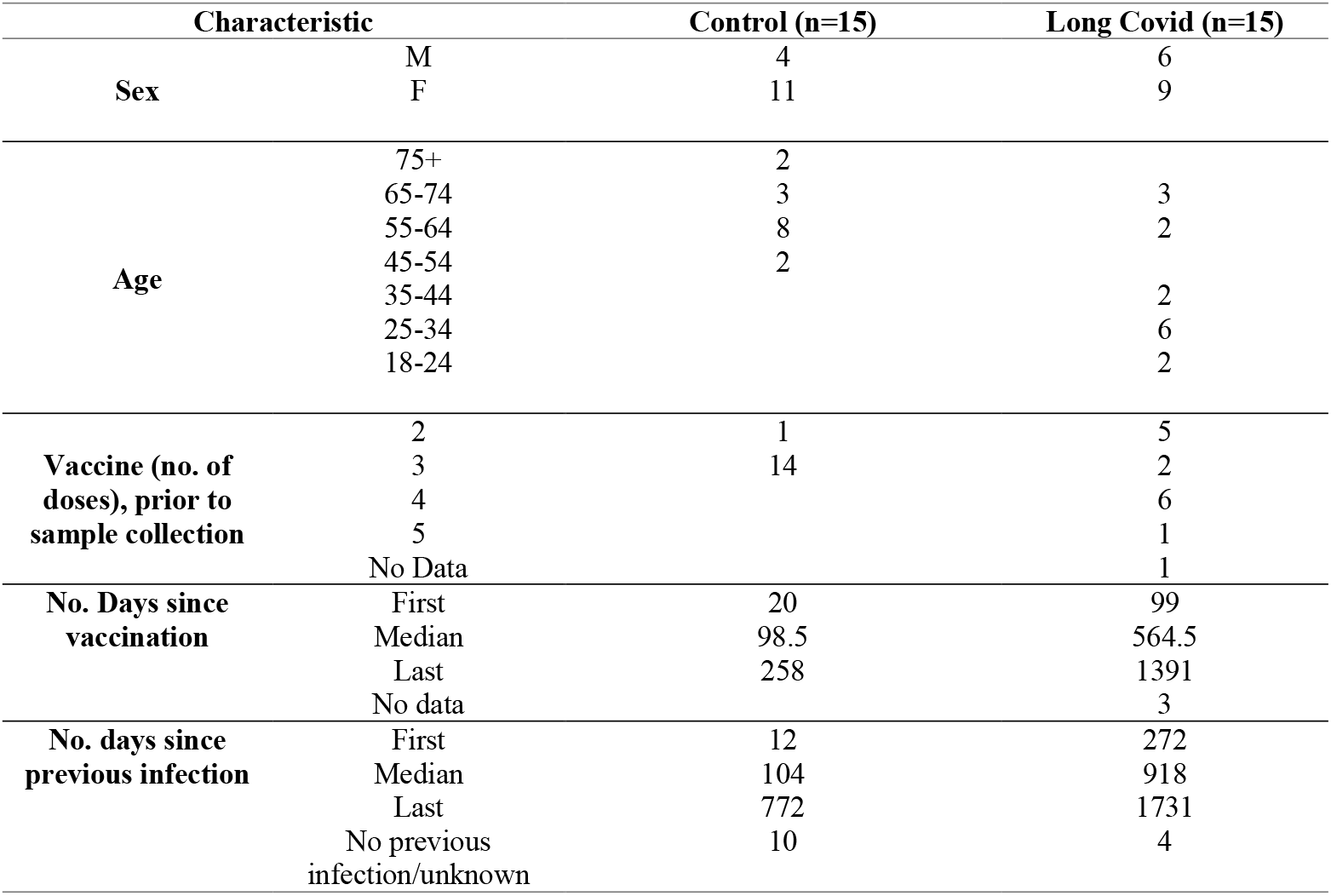
Patient Demographics.

| Characteristic |  | Control (n=15) | Long Covid (n=15) |
| --- | --- | --- | --- |
| Sex | M | 4 | 6 |
|  | F | 11 | 9 |
| Age | 75+ | 2 |  |
|  | 65-74 | 3 | 3 |
|  | 55-64 | 8 | 2 |
|  | 45-54 | 2 |  |
|  | 35-44 |  | 2 |
|  | 25-34 |  | 6 |
|  | 18-24 |  | 2 |
| Vaccine (no. of doses), prior to sample collection | 2 | 1 | 5 |
|  | 3 | 14 | 2 |
|  | 4 |  | 6 |
|  | 5 |  | 1 |
|  | No Data |  | 1 |
| No. Days since vaccination | First | 20 | 99 |
|  | Median | 98.5 | 564.5 |
|  | Last | 258 | 1391 |
|  | No data |  | 3 |
| No. days since previous infection | First | 12 | 272 |
|  | Median | 104 | 918 |
|  | Last | 772 | 1731 |
|  | No previous infection/unknown | 10 | 4 |

### Statistical analysis

Comparison of the concentrations of each IgG subclass between variants and cohorts was performed with a Shapiro-Wilks distribution normality and Mann-Whitney U tests were used for determining significance. Adjusted *p*-values were reported, corrected by the Holm-Šidák method to account for errors when using multiple comparisons. Mean-normalised distributions were calculated for each variant and merged for each patient in the CC and LC cohorts. The significance in the difference in the medians of on the resulting mean-normalised distributions were tested with Mann-Whitney U at the 95% confidence limit, again adjusted using Holm-Šidák method. For variance tests, significance was tested at the 95% confidence limits.

Correlation plots were constructed using the calculated concentrations and the summed concentrations for IgG in each variant, in each sample. All variants were treated together as a single dataset for the analysis of these data. The slopes of the lines of best fit were compared and tested for significance Fisher *r*-to-*z* transformation^38^ at the two-tailed 95% confidence limits. Distributions of the subclass percentage of the sum (IgG_x_/IgG_sum_*100) were plotted and analysed, with medians and IQR calculated.

A diagnostic classification was performed for the long COVID cohort using symmetric percentiles levels from the mean-normalised distributions for each variant with a variant above the upper of below the lower percentile providing a Boolean classification of long COVID. A diagnostic accuracy figure for the test was derived.

### Ethical Approval

The use of the Attomarker clinical samples was approved by the Bioscience Research Ethics Committee, University of Exeter, and each patient provided informed consent for their anonymised data to be used for COVID-related research.

## Results

The antibody concentration and quality were measured for each patient using the COVID Antibody Spectrum test to establish total IgG to each of the 14 variants and then repeated for each of the subclasses of IgG and IgE, are shown in Figure S1. The variant-dependent antibody subclass distributions from the control and long COVID cohorts are shown in Figure 1. These distributions identify healthy cohort reference ranges for each variant and antibody subclass, with differing inter-variant variability dependent on immunoglobulin subtype. Seventeen significant differences between medians of variants between the control and long COVID cohorts can be seen at the p < 0.05 and p <0.005 levels of significance, Figure 1, with the most significant differences observed for IgG4, then IgG3, then IgE.

**Figure 1.**
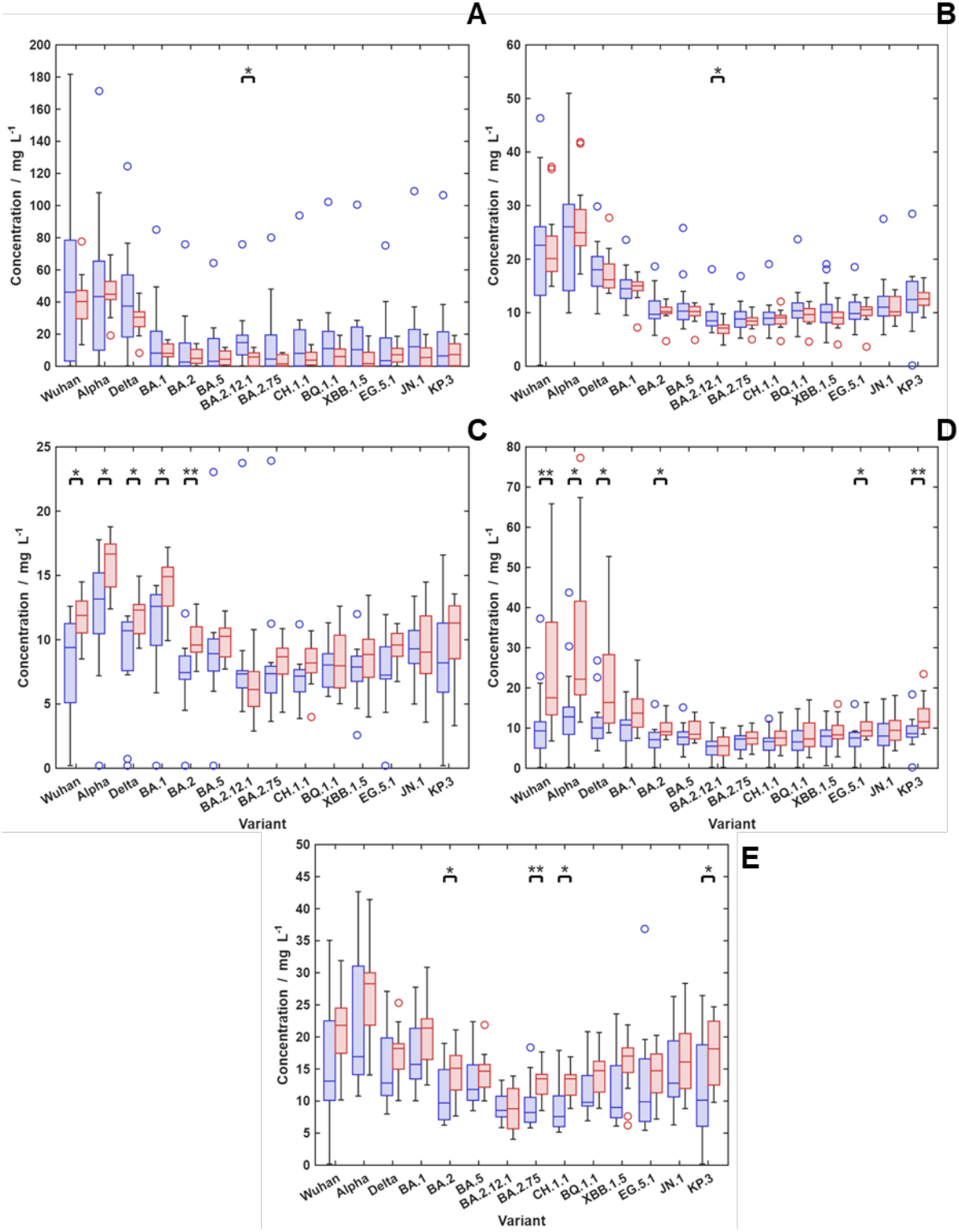
Antibody IgG subclass and IgE concentration distributions presented by SARS-CoV-2 Spike protein variant: CC (blue) and LC (red) cohorts. (A) IgG1; (B) IgG2, (C) IgG3, (D) IgG4 and (E) IgE. The boxplots show the medians and interquartile range. The vertical lines on each box point to the extremes of the distributions. Significant results to the p< 0.05 (*) and p<0.005 (**) level determined by the Mann-Whitney U test with Holm-Sidak correction applied

Inter-variant response comparisons can be made with mean-normalised concentration distributions for each variant calculated for the control and Long COVID cohorts, Figure S2. The median significance tests (using the Mann-Whitney U test) show the same seventeen differences, with the highest numbers of differences seen for IgG4, then IgG3 and, finally, IgE. The mean-normalised distributions for each variant can be merged, producing IgG subclass and IgE distributions for both cohorts, Figure 2.

**Figure 2.**
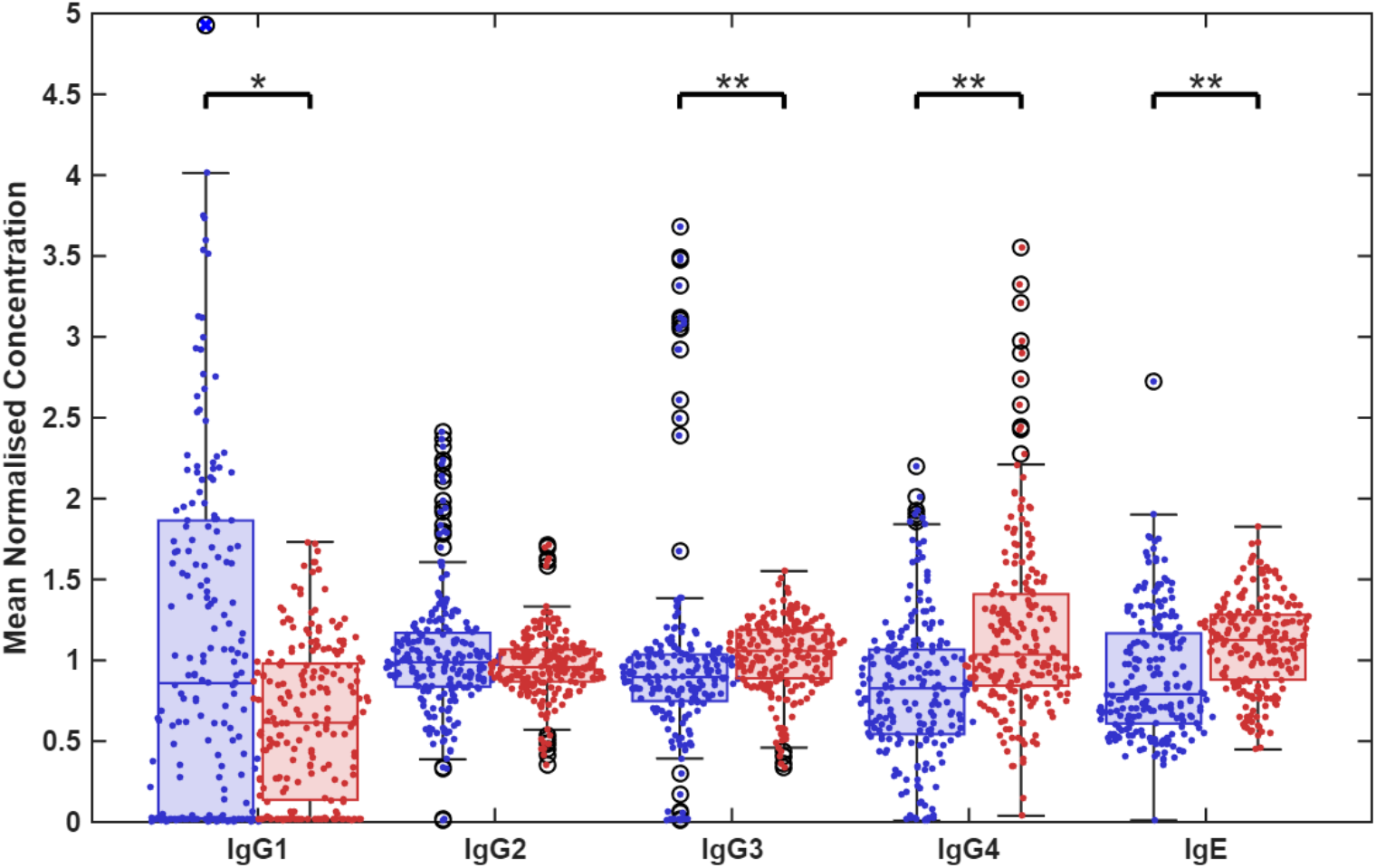
Mean-normalised concentration data for CC (blue) and LC (red). All variants are considered together for this analysis. Significant differences are present between the cohort medians for IgG1, IgG3 and IgG4 using a Mann Whitney U test with Holm-Sidak correction (Data shown in Table S2). (*) indicates significance p<0.05, (**) indicates significance p<0.005. Some outliers for the IgG1 control data lie outside of the plotted y-values and can be found in Table S3

The median significance tests for the IgG subtypes show significant differences: IgG1 is significantly lower (p < 0.05) in the long COVID cohort compared to controls; IgG2 shows no significant difference, whereas IgG3 (p < 0.005), IgG4 (p < 0.005), and IgE (p < 0.005) show significantly elevated levels in the long COVID cohort.

The correlation of IgG subclasses with summed IgG concentration are shown in Figure 3 for control and Long COVID cohorts. The control cohort shows a reasonable correlation for the production of IgG subclasses with total concentrations, Table 2, showing good correlation between IgG1 and total IgG in controls and long COVID patients. Critically, the order of gradients of the correlations different in Figure 3 A and B; IgG1-IgG2-IgG3-IgG4-IgE for the controls (highest to lowest) and for long COVID patients IgG1-IgG4-IgG2-IgE-IgG3 (highest to lowest); those with long COVID are more likely to produce IgG4 and IgE, whilst IgG2 is unchanged. IgG3 shows a higher median, but below the upper outliers in the control group, Figure 2.

**Table 2.** The linear fit data for IgG subclasses against the total IgG for the same sampleshown in Figure 3. A Fisher r-to-z transformation was used to test the significance of differences between the CC and LC cohorts, calculating a two-tailed p-value.

| Subclass | Control Cohort |  |  | Long Covid Cohort |  |  | Fisher r-to-z<br>Two-tailed p-value<br>(n = 210<br>observations) | Fisher r-to-z<br>Two-tailed p-value<br>(n = 15<br>samples) |
| --- | --- | --- | --- | --- | --- | --- | --- | --- |
|  | m | c | r<sup>2</sup> | m | c | r<sup>2</sup> |  |  |
| IgG1 | 0.66 | -12.9 | 0.95 | 0.45 | -8.9 | 0.85 | $1.1 \times 10^{-9}$ | 0.14 |
| IgG2 | 0.14 | 6.0 | 0.72 | 0.18 | 3.7 | 0.87 | $8.3 \times 10^{-6}$ | 0.28 |
| IgG3 | 0.12 | 3.1 | 0.67 | 0.07 | 6.7 | 0.52 | $1.8 \times 10^{-2}$ | 0.57 |
| IgG4 | 0.09 | 3.8 | 0.47 | 0.30 | -1.5 | 0.73 | $1.6 \times 10^{-5}$ | 0.30 |
| IgE | 0.03 | 11.43 | 0.05 | 0.10 | 11.13 | 0.32 | $3.5 \times 10^{-3}$ | 0.48 |

**Figure 3.**
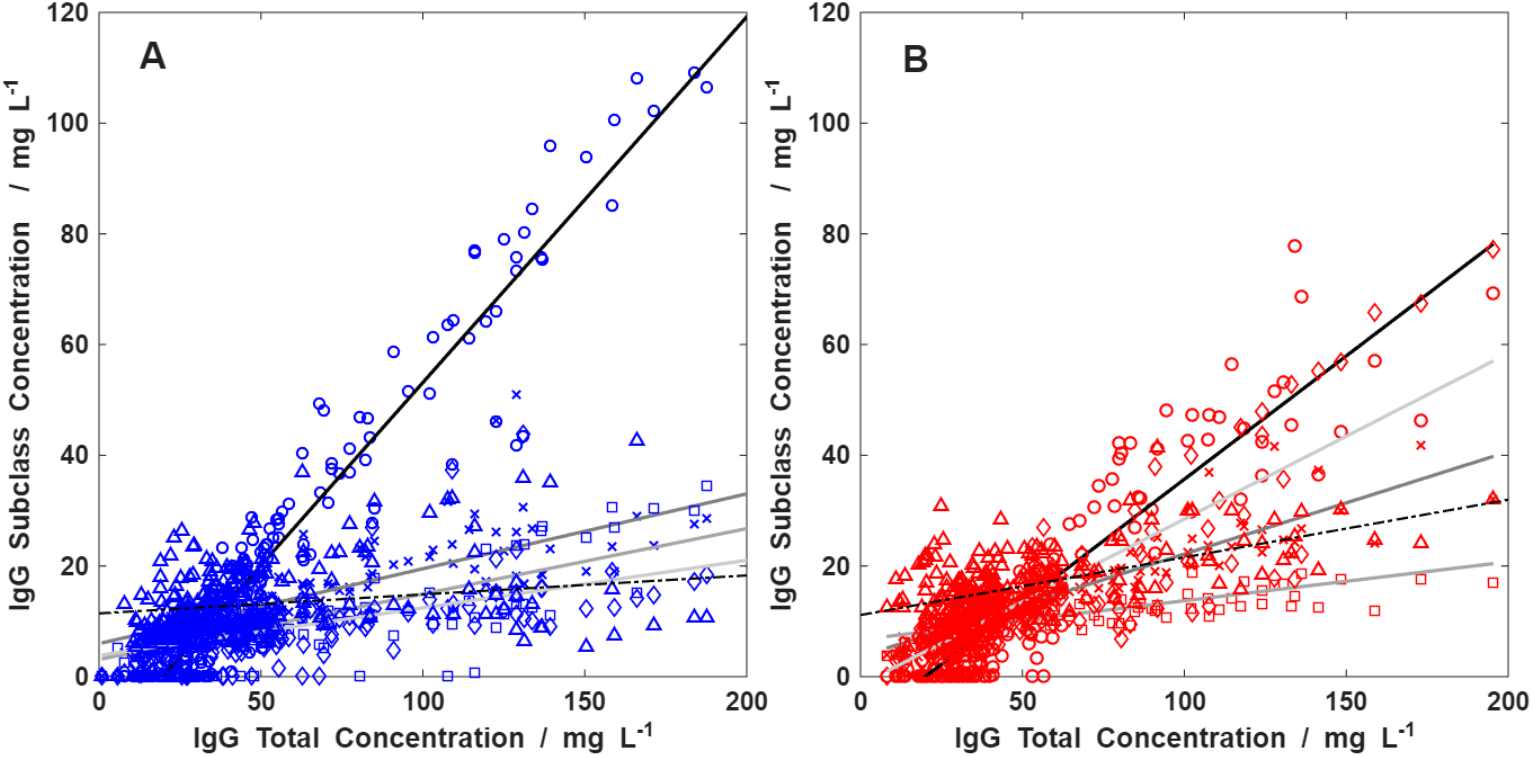
Correlation between Total IgG and subclass measurements: A) Control Cohort, B) Long Covid Cohort. Marker shape denotes subclass: IgG1 (o), IgG2 (x), IgG3 (□), IgG4 (◊) and IgE (Δ). Regression lines are ordered vertically and IgG1 and IgG4 production dominates in long COVID patients compared with IgG1 and IgG2 in controls; A) controls IgG1-IgG2-IgG3-IgG4-IgE; and B) long COVID IgG1-IgG4-IgG2-IgE-IgG3.

The significance of the correlation in small samples for the correlation coefficients *r*^2^ when tested by number variant tests performed (*n*=210) and the total number of different patient samples (*n*=15) was tested using the Fisher *r*-to-*z* transformation^38^ for all total observations by variant, and for the 15 samples. All correlation coefficients are significant when compared for the full 210 multivariant tests, but none when considering the 15 patient samples alone, Table 2.

The mean-normalised distributions for each of the variants were tested for diagnostic accuracy for both cohorts. A symmetric reference interval from the control cohort provides an upper and lower threshold; any variant above or below the control destitution threshold is considered in long COVID cohort as a Boolean operator. The best diagnostic sensitivity and specificity derived from the distribution percentiles classification was calculated for the long COVID cohort for each antibody subclass. The highest diagnostic accuracy of 80% was derived from the 60^th^ symmetric percentile for IgG3, Figure 3 and Table 3; the next highest was IgG4.

**Table 3.** The diagnostic accuracy shown for the long COVID classification based on threshold derived from symmetric reference range percentiles for the antibody subclass IgG1, IgG3, IgG4 and IgE giving sensitivity, specificity and accuracy.

| Metric / Concentration | Reference<br>Interval / % | Sensitivity / % | Specificity / % | Accuracy / % |
| --- | --- | --- | --- | --- |
| **IgG1 < Reference** | 55 | 66.7 | 46.7 | 56.7 |
| **IgG3 > Reference** | 60 | 80.0 | 80.0 | 80.0 |
| **IgG4 > Reference** | 65 | 73.3 | 73.3 | 73.3 |
| **IgE > Reference** | 65 | 73.3 | 60.0 | 66.7 |

The reference interval can be used to provide immunotype classifications, assessing particular variant dependence outside the symmetric interval, as shown in Figure 4; IgG1 variants only produced subthreshold classifications, hypoimmune, and IgG3, IgG4 and IgE produced above threshold classifications, hyperimmune, consistent with differences seen in Figure 2. The later variants in IgG1 are contributing to the long COVID classification hypoimmune, below the threshold, whereas all other subclasses are hyperimmune, above the threshold.

**Figure 4.**
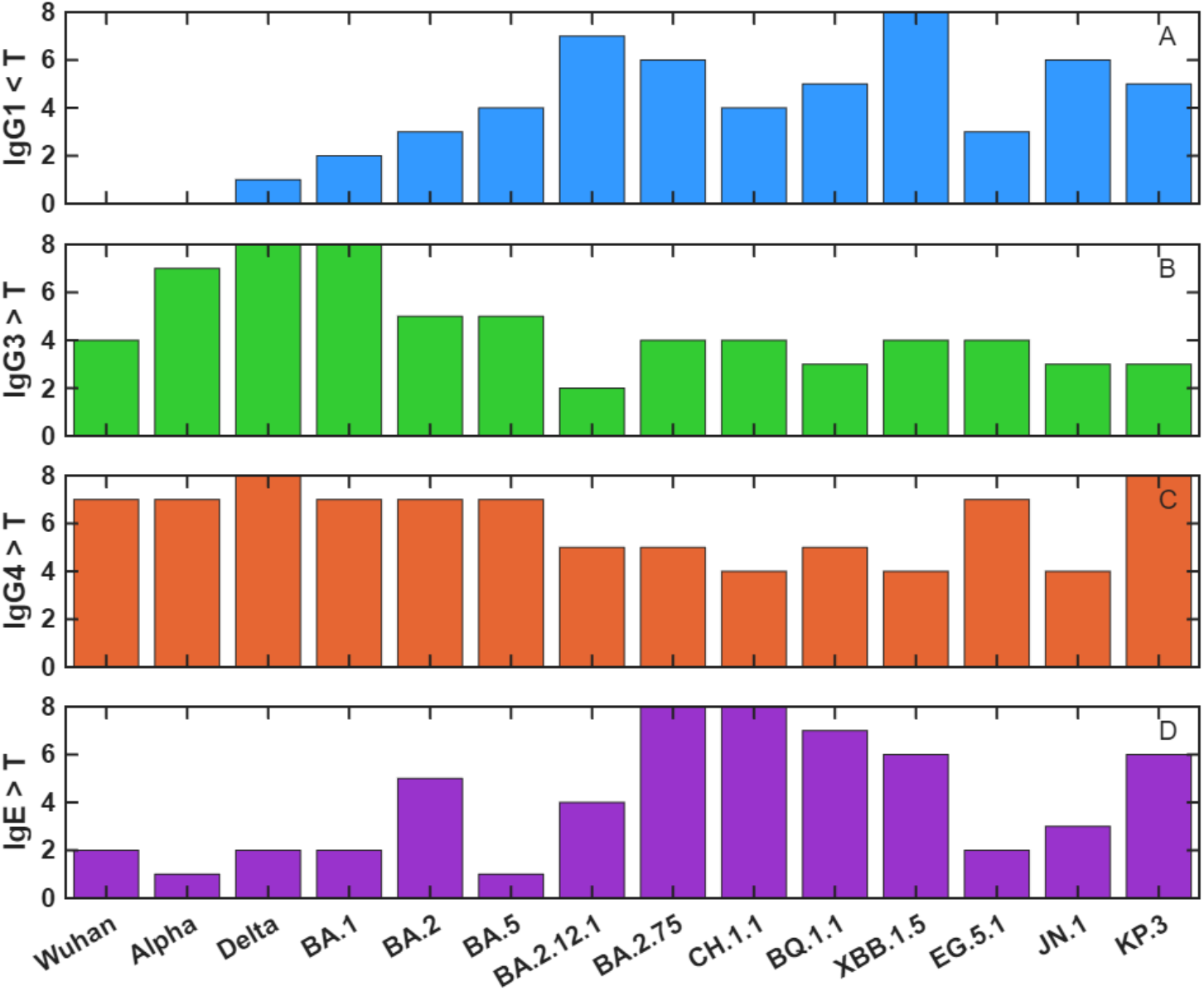
The variant profiles showing the long COVID classification triggers: (A) IgG1 below threshold, T; and (B) IgG3 (C) IgG4 (D) IgE above threshold, T, derived from the percentile (T) in the mean-normalised variant concentration distributions in the Control Cohort.

## Discussion

The function of antibody subclasses in the systemic response to antigens^39^ provides more understanding of the immune system response beyond a marker of previous exposure to an infection. Subclass characteristics report on the efficacy of the vaccine response with the presence of high quality IgG1 and IgG3 subclasses and elevation of IgG4 and IgE suggestive of chronic conditions^40^. Differential antibody subclass production is a measure of the inflammatory state of the immune system and IgG4, in particular, is associated with IgG4-Related Disease^8,41^ (IgG4-RD). Many symptoms of IgG4-RD have been reported for long COVID such as fibroinflammatory lesions over multiple organs as well as chronic allergic disorders, autoimmune disease and chronic infections such as hepatitis^42^. Similarly, elevated IgE to specific antigens is associated with food allergies, hyper-IgE syndrome^43^, autoimmune disease such as urticaria, and other persistent infections. Much of the underlying pathophysiology of long COVID is poorly understood but has been characterised by fibrosis and inflammation^38^. Elevated levels of IgG4 and IgE in long COVID patients is consistent with chronic inflammation, and for IgE points to one mechanism for mast cell activation^44^. A profile of the IgG subclasses and IgE to the spike protein antigen may, therefore, have potential for the differential classification of long COVID patients and a potential role in companion diagnostics for possible therapeutic interventions.

In this study, the levels of IgG subclasses and IgE have been measured in two cohorts; samples from non-long COVID controls and those classified with having long COVID, for a panel of variants spanning the pandemic. The subclass levels have been quantified for total IgG and IgG1 because of the availability of accurate standard materials. However, similar standard materials for IgG2-4 and IgE were not readily available. The detection materials used in the assay are all monoclonal antibodies and so inter-class comparisons can be made, as shown in Figure 2. The mean-normalisation implicitly assumes that differences between the variants scale linearly with the mean, which is implicit in the mass-sensitivity of the biophotonic technique. There remain some sources of bias from mean-normalisation: large outliers can skew the mean, especially in small cohorts and can compress distributions. However, as determined from the significance testing, Table S2, the variation between all variant concentrations and all subclasses (including IgE), 24% of all possible comparisons is reproduced in the mean-normalised analysis. Combining the distributions for the inter-class comparisons shows increased IgG3 and IgG4, and decreased IgG1, in relation to the long COVID cohort compared to the controls. IgG2 levels are similar in both cohorts, likely because IgG2 is associated with bacterial infection^45,46^ and long COVID is typically associated with a SARS-CoV-2 viral infection.

The IgE concentrations measured in both cohorts are extremely high, in the range 10-20 mg/L which may be compared with the severe peanut allergy levels of 667 μg/L^47^. The absolute concentrations are over-estimated in this study as the detection antibody is a polyclonal, potentially binding in multiple locations on the spike protein surface. The over-estimate is a stearic-interaction limited by the proximity of binding sites on the spike protein suggesting a four- to ten-fold overestimate in the cencentration. Notwithstanding the overestimate, elevated IgE to spike protein in the control cohort, and so the general population, appears to be high, suggesting rigorous standardisation and sampling must be considered to characterise the risk. This elevated (hyperimmune) response has potential to manifest in the wider population as unexpected medical presentations and adverse reactions. The over-estimate transfers consistently between both sample cohorts allowing them to be compared directly: there are significantly elevated IgE levels for the long COVID cohort compared with the controls, Figure 2. This suggests a mechanism for mast cell activation *via* an IgE-mediated response^44^, and supports previous findings showing histamine and cytokine release following a SARS-CoV-2 infection^48^. Further, the IgE affinity may also be important as a companion diagnostic^49^.

A preliminary diagnostic analysis has been performed on the long COVID cohort using the self-declaration that a patient has long COVID as agreed by the physician (and the WHO classification) as the dichotomous marker. The upper and lower percentile limits in the distribution of controls were varied symmetrically in the mean-normalised concentration distributions. A long COVID Boolean classification was assigned if the patient had an antibody below the lower percentile or above the upper percentile for any variant. The sensitivity, specificity and accuracy are calculated for each percentile, Table 3, with an 80% accuracy for elevated IgG3 with IgG4 and IgE slightly lower in accuracy. The classification allows for three types of long COVID patient, IgG3 antibody deficient, hypo-immune, elevated IgG4 or IgE hyperimmune, and those patients for whom there is no deviation from the reference range. Given the unknown accuracy of the long COVID classification as post-viral complications for more than 20 weeks, the result is promising and potentially mechanistic associated with persistence of virus or antigen and poor concentrations of IgG1 antibodies to a particular variant.

The detailed variant analysis shows that the variant responsible for the hypo- or hyper-immune classification is shown in distributions, Figure 4, shows that IgG1 hypo-immunity does not occur for the early variants, consistent with efficient vaccination and good quantity of IgG1 antibodies in the vaccine response; later variants show gaps consistent with vaccine antibody optimisation to epitopes on the spike protein that no longer exist in later variants. The hyperimmune variant distributions for IgG3 and IgG4, Figure 4B and C, are similar, with most elevation occurring to the early variants; conversely IgE is most elevated for later variants, Figure 4D.

Long COVID patients show some interesting, potentially diagnostic, IgG subclass distributions^50^, Figure 3, which may reflect the underlying pathophysiology and potential new interventions. The two most frequently postulated pathophysiologies^33^ for long COVID are persistent virus and autoimmunity. Persistent virus with intermittent recurrence of infection could lead to chronic inflammation and IgG1, IgG3 and IgG4 production and autoantibody subclass distributions such as in rheumatoid arthritis^51^ show elevated IgG1 and IgG4. IgG3 was observed in LC here which has been seen in patients with a positive SARS-CoV-2 infection ^52,53^. There are also reports of class switching to IgG4 following mRNA vaccination and prior inflammation history^18^ and long COVID patients have been shown to have differential IgG4/IgG1 ratio following multiple vaccination compared with controls^54,55^.

IgG4 is often considered a useful marker for determining clinical outcomes^49^ and the multivariant IgG subclass analysis suggests a number of possible new interventions for long COVID from disease states with elevated IgG4 targeting immune modulation^56^. B cell depletion (inebilizumab, rituximab, obexelimab), signal inhibition (BTK inhibitors such as rilzabrutinib) IgG clearance (FcRn inhibitors such as efgartigimod) and cytokine blockades (such as IL-4/IL-13 inhibitor, dupilumab) have been used as therapies for IgG4-RD and IgG4 autoimmune diseases.

## Conclusions

The multivariant IgG subclass analysis for long COVID patients has produced some novel insights into patterns of adaptive response, chronic inflammation, chronic exposure to antigens and the production of IgG4 and IgE. The persistent virus signatures in the antibody subclasses have been seen in several other conditions, and in particular, IgG4-related disease is reported with other persistent viruses such as Epstein-Barr virus and HIV-1^57,58^. The autoimmune subclass distributions similarly seen in rheumatoid arthritis^51^ points to additional diagnostic signatures suggesting that both prevalent theories for long COVID pathophysiology would also have a signature in the IgG subclass variant spectrum. Considering the small sample subset herein producing a diagnostic accuracy for long COVID at 80%, suggests there may be routes to diagnosis and companion diagnostics in the antibody profile of the adaptive immune response.

## Data Availability

Available on request

## Funding

The project was funded by Attomarker Ltd, the alumni of the University of Exeter donating to the COVID appeal in Prof Shaw’s group, and the patients taking the COVID Antibody Spectrum Test. Abishek Elangovan worked on the study as part of their undergraduate studies at the University of Exeter.

## Declaration of Interests

Prof Shaw is the Founder, CEO and Director of Attomarker Ltd, a spin-out company from his research group at the University of Exeter, which is also a shareholder.

## Supplementary Data

**Figure S1.**
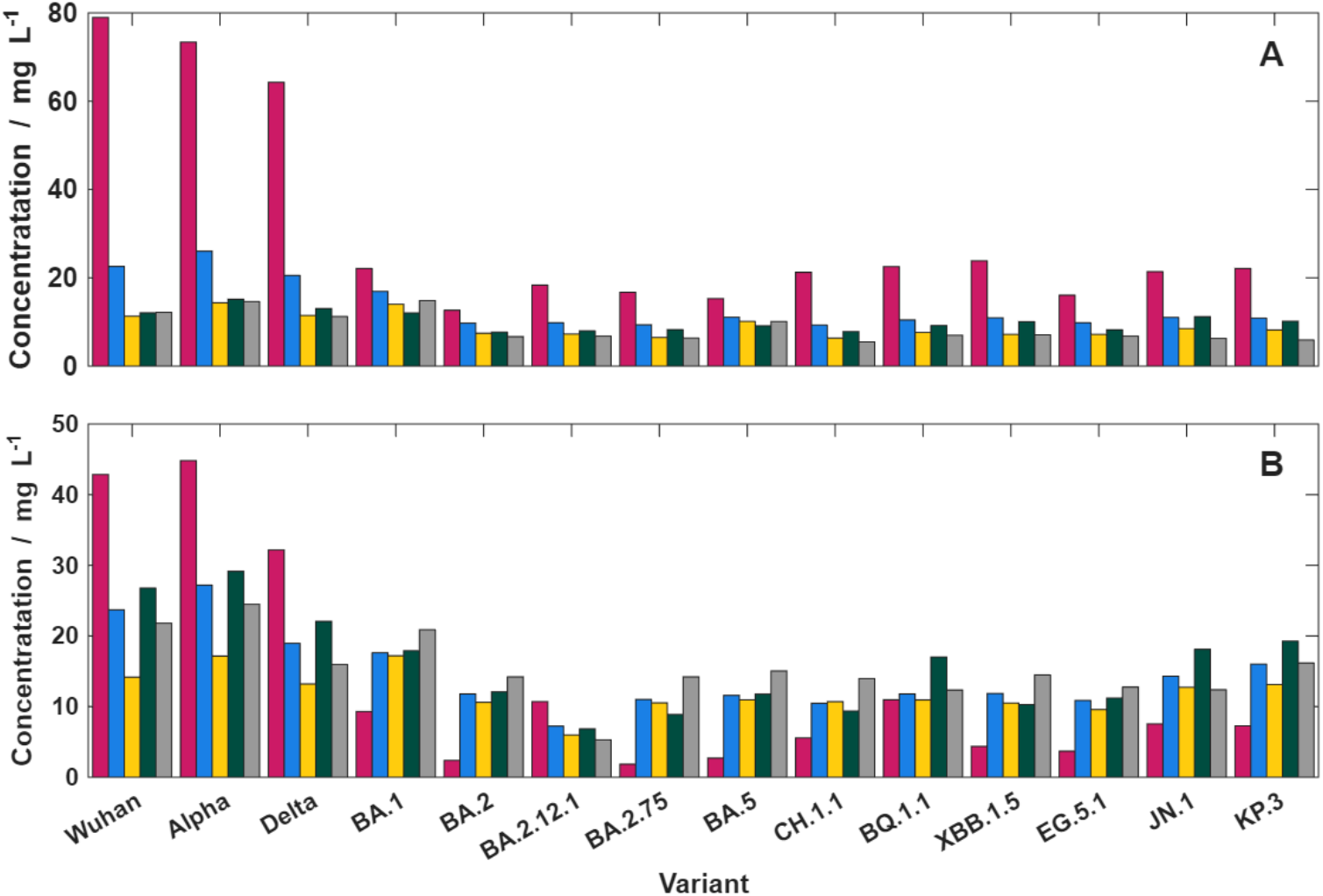
Single patient results showing the variant specific response to each IgG subclass measured separately for IgG1-IgE but for all variants simultaneously: A) Control sample and B) Long Covid sample: IgG Subclass concentrations IgG1-4: coloured as pink (IgG1), blue (IgG2), yellow (IgG3), green (IgG4) and grey (IgE).

**Table S1.** Materials table of detection antibodies. Running concentrations for all detection reagents are diluted using RB1.

| Detection antibody | Manufacturer | Product Code |
| --- | --- | --- |
| Monoclonal Mouse Anti-Human IgG3 | Biorad | 5247-9850 |
| Monoclonal Mouse Anti-Human IgG2 | Biorad | 5224-9850 |
| Monoclonal Mouse Anti-Human IgG4 | Biorad | 919001 |
| Monoclonal Mouse Anti-Human IgG1 | Biorad | 5218-9850 |
| Polyclonal Goat Anti-Human IgE | Biorad | STAR147 |
| Anti-Human CRP | Randox | CP7950 |

**Figure S2.**
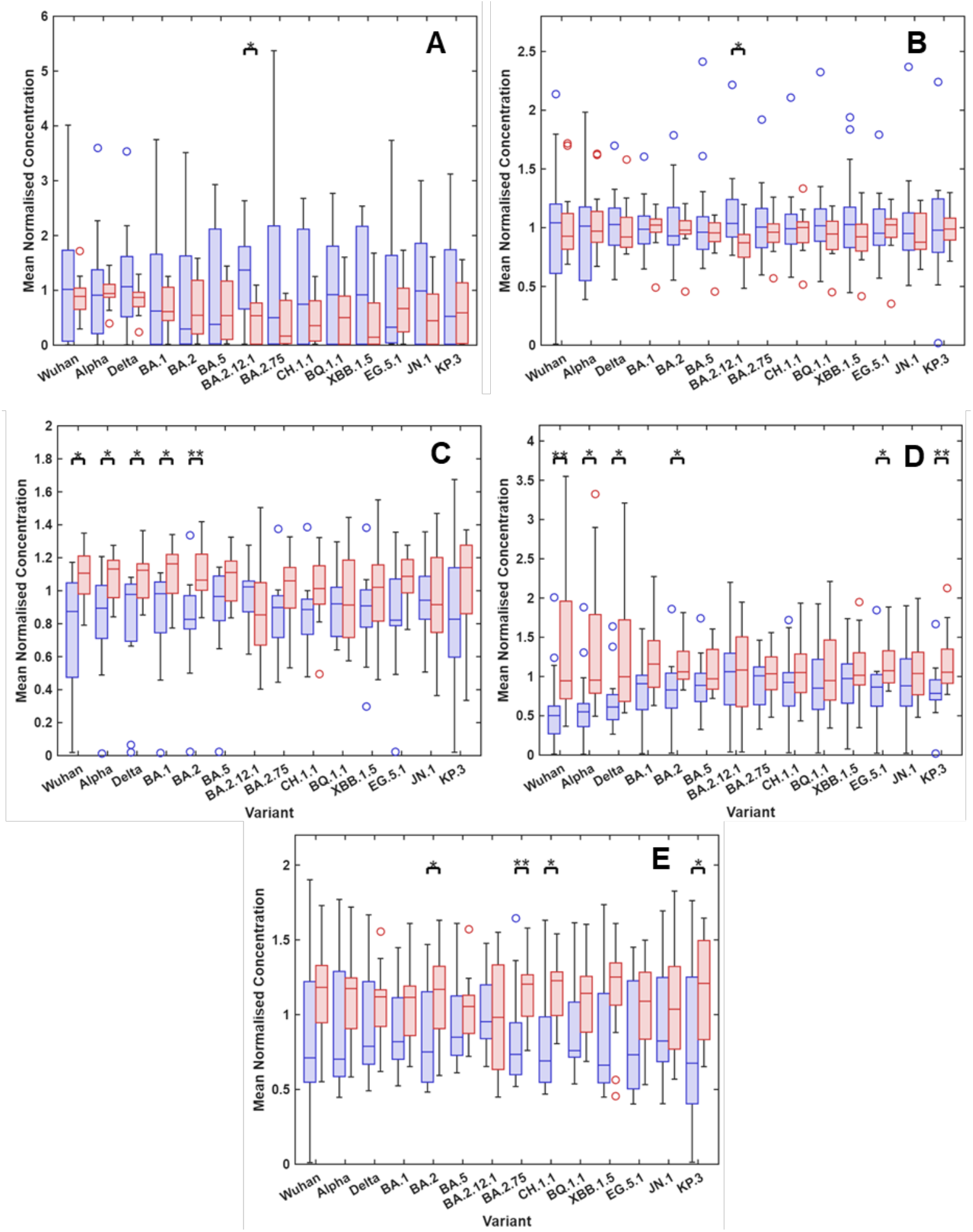
Mean-normalised concentration data presented by variant for the CC (blue) and LC (red) cohorts. Significant results to the p < 0.05 (*) and p < 0.005 (**) level determined by the Mann-Whitney U test with Holm-Sidak correction applied.

**Table S2.** Holm-Sidak Adjusted Significance values in median differences comparing the mean normalised concentration data of the CC and LC groups. ‘All’ denotes where all variant data was combined and tested together.

| Channel | IgG1 | IgG2 | IgG3 | IgG4 | IgE |
| --- | --- | --- | --- | --- | --- |
| All | 0.005 | 0.846 | 0.000 | 0.000 | 0.000 |
| Wuhan | 0.968 | 0.994 | 0.035 | 0.003 | 0.409 |
| Alpha | 0.998 | 0.815 | 0.026 | 0.006 | 0.481 |
| Delta | 0.490 | 0.876 | 0.032 | 0.013 | 0.088 |
| BA.1 | 1.000 | 0.538 | 0.019 | 0.080 | 0.185 |
| BA.2 | 1.000 | 0.685 | 0.003 | 0.010 | 0.040 |
| BA2.12.1 | 0.007 | 0.004 | 0.510 | 1.000 | 1.000 |
| BA2.75 | 0.362 | 0.977 | 0.139 | 0.574 | 0.004 |
| BA.5 | 0.927 | 1.000 | 0.100 | 0.200 | 0.731 |
| CH1.1 | 0.393 | 0.999 | 0.053 | 0.248 | 0.006 |
| BQ1.1 | 0.301 | 0.384 | 1.000 | 0.981 | 0.053 |
| XBB1.5 | 0.087 | 0.228 | 0.834 | 0.767 | 0.065 |
| EG.5.1 | 1.000 | 1.000 | 0.066 | 0.018 | 0.267 |
| JN.1 | 0.243 | 0.944 | 1.000 | 0.877 | 0.957 |
| KP.3 | 0.828 | 0.723 | 0.269 | 0.003 | 0.044 |

**Table S3.**
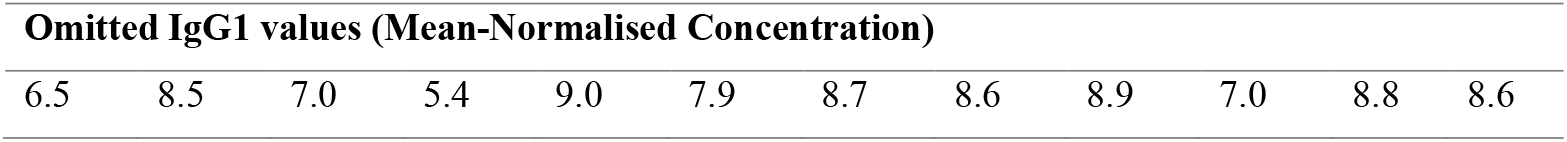
Omitted data points (above cutoff) from Figure 2.

**Table S4.**
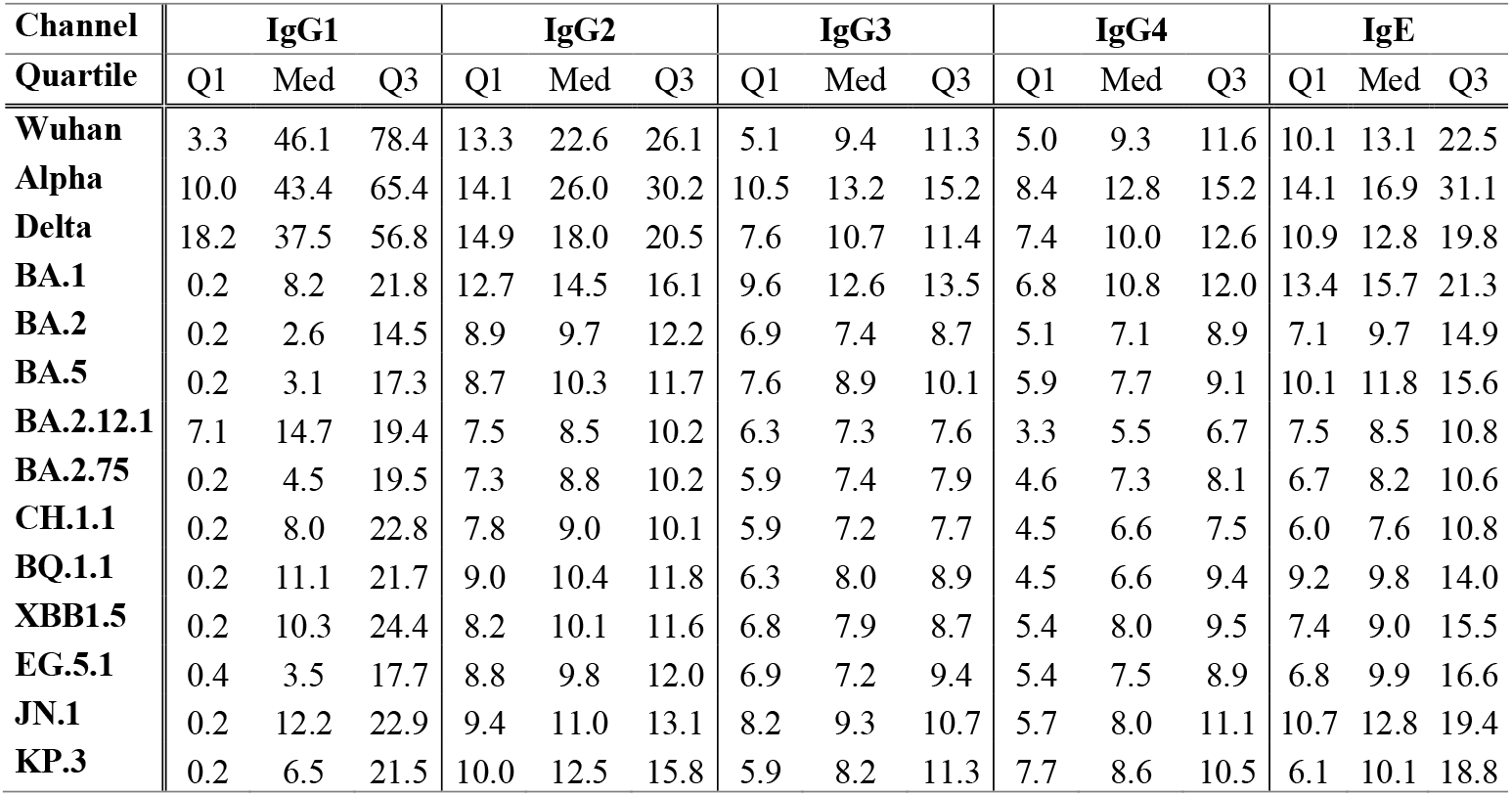
Quartiles of Control Sample Concentrations.

| Channel | IgG1 |  |  | IgG2 |  |  | IgG3 |  |  | IgG4 |  |  | IgE |  |  |
| --- | --- | --- | --- | --- | --- | --- | --- | --- | --- | --- | --- | --- | --- | --- | --- |
|  | Q1 | Med | Q3 | Q1 | Med | Q3 | Q1 | Med | Q3 | Q1 | Med | Q3 | Q1 | Med | Q3 |
| Wuhan | 3.3 | 46.1 | 78.4 | 13.3 | 22.6 | 26.1 | 5.1 | 9.4 | 11.3 | 5.0 | 9.3 | 11.6 | 10.1 | 13.1 | 22.5 |
| Alpha | 10.0 | 43.4 | 65.4 | 14.1 | 26.0 | 30.2 | 10.5 | 13.2 | 15.2 | 8.4 | 12.8 | 15.2 | 14.1 | 16.9 | 31.1 |
| Delta | 18.2 | 37.5 | 56.8 | 14.9 | 18.0 | 20.5 | 7.6 | 10.7 | 11.4 | 7.4 | 10.0 | 12.6 | 10.9 | 12.8 | 19.8 |
| BA.1 | 0.2 | 8.2 | 21.8 | 12.7 | 14.5 | 16.1 | 9.6 | 12.6 | 13.5 | 6.8 | 10.8 | 12.0 | 13.4 | 15.7 | 21.3 |
| BA.2 | 0.2 | 2.6 | 14.5 | 8.9 | 9.7 | 12.2 | 6.9 | 7.4 | 8.7 | 5.1 | 7.1 | 8.9 | 7.1 | 9.7 | 14.9 |
| BA.5 | 0.2 | 3.1 | 17.3 | 8.7 | 10.3 | 11.7 | 7.6 | 8.9 | 10.1 | 5.9 | 7.7 | 9.1 | 10.1 | 11.8 | 15.6 |
| BA2.12.1 | 7.1 | 14.7 | 19.4 | 7.5 | 8.5 | 10.2 | 6.3 | 7.3 | 7.6 | 3.3 | 5.5 | 6.7 | 7.5 | 8.5 | 10.8 |
| BA2.75 | 0.2 | 4.5 | 19.5 | 7.3 | 8.8 | 10.2 | 5.9 | 7.4 | 7.9 | 4.6 | 7.3 | 8.1 | 6.7 | 8.2 | 10.6 |
| CH1.1 | 0.2 | 8.0 | 22.8 | 7.8 | 9.0 | 10.1 | 5.9 | 7.2 | 7.7 | 4.5 | 6.6 | 7.5 | 6.0 | 7.6 | 10.8 |
| BQ1.1 | 0.2 | 11.1 | 21.7 | 9.0 | 10.4 | 11.8 | 6.3 | 8.0 | 8.9 | 4.5 | 6.6 | 9.4 | 9.2 | 9.8 | 14.0 |
| XBB1.5 | 0.2 | 10.3 | 24.4 | 8.2 | 10.1 | 11.6 | 6.8 | 7.9 | 8.7 | 5.4 | 8.0 | 9.5 | 7.4 | 9.0 | 15.5 |
| EG.5.1 | 0.4 | 3.5 | 17.7 | 8.8 | 9.8 | 12.0 | 6.9 | 7.2 | 9.4 | 5.4 | 7.5 | 8.9 | 6.8 | 9.9 | 16.6 |
| JN.1 | 0.2 | 12.2 | 22.9 | 9.4 | 11.0 | 13.1 | 8.2 | 9.3 | 10.7 | 5.7 | 8.0 | 11.1 | 10.7 | 12.8 | 19.4 |
| KP.3 | 0.2 | 6.5 | 21.5 | 10.0 | 12.5 | 15.8 | 5.9 | 8.2 | 11.3 | 7.7 | 8.6 | 10.5 | 6.1 | 10.1 | 18.8 |

## Notes

### Competing Interest Statement

Prof Shaw is founder and CEO of Attomarker Ltd, a spin-out from the University of Exeter, which is also a shareholder.

### Funding Statement

University of Exeter Alumni as a donation to Prof Shaw's reproach group and Attomaker Ltd.

